# Pooling of samples to optimize SARS-CoV-2 diagnosis by RT-qPCR: comparative analysis of two protocols

**DOI:** 10.1101/2020.08.24.20181008

**Authors:** Fabiana Volpato, Daiana Lima-Morales, Priscila Lamb Wink, Julia Willig, Fernanda Paris, Patricia Ashton-Prolla, Afonso Luís Barth

## Abstract

RT-qPCR for SARS-CoV-2 is the main diagnostic test used to identify the novel coronavirus. Several countries have used large scale SARS-CoV-2 RT-qPCR testing as one of the important strategies for combating the pandemic. In order to process the massive needs for coronavirus testing, the usual throughput of routine clinical laboratories has reached and often surpassed its limits and new approaches to cope with this challenge must be developed. This study has aimed to evaluate the use pool of samples as a strategy to optimize the diagnostic of SARS-CoV-2 by RT-qPCR in a general population. A total of 220 naso/orofaryngeal swab samples were collected and tested using two different protocols of sample pooling. In the first protocol (Protocol A); 10 clinical samples were pooled before RNA extraction. The second protocol (Protocol B) consisted of pooling the already extracted RNAs from 10 individual samples. Results from Protocol A were identical (100% agreement) with the individual results. However, for results from Protocol B, reduced agreement (91%) was observed in relation to results obtained by individual testing. Inconsistencies observed were related to RT-qPCR results with higher Cycle Thresholds (Ct > 32.73). Furthermore, in pools containing more than one positive individual, the Ct of the pool was equivalent to the lowest Ct among the individual results. These results provide additional evidence in favor of the clinical use of pooled samples for SARS-CoV-2 diagnosis by RT-qPCR and suggest that pooling of samples before RNA extraction is preferrable in terms of diagnostic yield.

## Introduction

Due to the exponential increase of respiratory syndromes by SARS-CoV-2, the World Health Organization (WHO) declared COVID-19 a pandemic of worldwide importance (1). The rapid propagation of the virus significantly increases the demand on the health care system. In response to the challenge of identifying and isolating infected patients, in an attempt to minimize spread of the disease, two different types of tests have been used: real time Reverse Transcriptase Polymerase Chain Reaction (RT-qPCR) for SARS-CoV-2 diagnosis mainly in upper respiratory tract samples and serum IgM and/or IgG antibody detection assays to detect past infection and possibly immunity to the disease.

In order to avoid diagnostic cross-reactivity with other endemic coronaviruses, most RT-qPCR assays include primers for at least two different molecular targets, such as nucleocapsid proteins 1 (N1) and 2 (N2) (2). A major advantage of real time assays is that amplification and analysis of the final product are performed simultaneously in a closed system. This procedure minimizes false-positive results associated with amplification of unspecific products (2).

Several countries have used RT-qPCR in large scale efforts as a mainstream initiative for combating the SARS-CoV-2 pandemic. To enable massive coronavirus testing thousands of laboratories worldwide have expanded their routine and in several countries high-troughput automated testing was implemented mainly in reference centers. However, many laboratories, especially in underserved areas became overloaded and new approaches to cope with this ever increasing and long lasting challenge are needed, especially in developing countries and if a strategy of massive testing, including asymptomatic individuals is adopted. A few studies have provided encouraging results that support the use of sample pooling as a robust and cost/time-saving approach in this extreme scenario. In this brief report, we provide additional evidence in favor of SARS-CoV-2 RT-qPCR analysis with pooled samples using two different methodologies.

## Materials and Methods

### Sample Pooling

A total of 220 samples from different patients were obtained by oro/nasopharyngeal swabbing performed by trained personnel in a single institution. Individual swabs were mixed with 3mL of NaCl 0.9% solution. Two strategies of pooling samples were evaluated: 1) Protocol A: consisted of 10 samples grouped (100μL from each sample) before RNA extraction; 2) Protocol B: consisted of 10 samples group (4μL of each) after the individual RNA extraction. Results from both protocols were compared to individual analyses of the same samples, performed in parallel to the analysis of the pools. RNA was extracted from all samples using the Abbott m2000sp automated system (Abbott Laboratories, Chicago, USA) and the Abbott Sample Preparation System – 4x24 Preps (Promega Corporation, Wisconsin, USA).

#### RT-qPCR Reaction

A previously validated SARS-CoV-2 RT-qPCR protocol following the CDC guidelines and primers was used. In brief, Superscript III (SSIII) one step RT-qPCR system (Thermo Fisher Scientific Inc, California, USA) was used for RT-qPCR reactions. The master mix was composed of 5uL of 2X reaction buffer (0.4 mM of each dNTP and 6 mM MgSO_4_); 0.2μL of SuperScript™ III RT/Platinum™ Taq Mix; 0.2μL of ROX (dilution 1:10); 0.75μL of combined primers/probes mix of nCOV1 (N1 primer) or nCOV2 (N2 primer) or RP (2019-nCoV RUO Kit, IDT, Integrated DNA Technologies Inc, Iowa, USA) and 4μL of extracted RNA. The human ribonuclease P gene (RNAse P) was used as internal control to monitor nucleic acid extraction, specimen quality and presence of reaction inhibitors. Thermal cycling was performed at 50°C for 30 min for reverse transcription, followed by 95 °C for 2 min and then 45 cycles of 95°C for 15s, 55 °C for 35s in QuantStudio® 3 Applied Biosystems™ (Applied Biosystems, Massachusetts, USA). The result was considered “negative” when neither N1 nor N2 targets amplified and “positive” when both N1 and N2 targets amplified. RT-qPCR was considered “inconclusive” when only one target (N1 or N2) amplified. Individual samples and pools with positive results had the values of the Cycle Threshold (Ct) recorded. In order to analyze performance of the assays with pooled samples, Ct values of individual and pooled sample runs were compared.

To minimize inter-experiment variability (“batch effects”) simultaneous analysis of the individual samples and their corresponding pools was undertaken in the same RT-qPCR reaction. The 220 samples included in this study were sorted in 22 pools with 10 clinical samples each, for each protocol. The individual composition of the 22 pools for each protocol (A and B) was as follows: all negative samples (1/22), one inconclusive sample (1/22), only one positive sample (8/22), two positive samples (7/22), three positive samples (3/22), four positive samples (1/22), and five positive samples (1/22) (Table 1)

**Table 1:** Cycle Threshold results of pooling samples for detection of 208 SARS-CoV-2 by RT-qPCR.

| Pool Identification | Protocol A* Ct N1 | Protocol A Ct N2 | Protocol B** Ct N1 | Protocol B Ct N2 | Number of positive samples in the pool | Value of lower Ct positive in the pool N1 | Value of lower Ct positive in the pool N2 |
| --- | --- | --- | --- | --- | --- | --- | --- |
| POOL 1 | Negative | Negative | Negative | Negative | 0 | - | - |
| POOL 2 | 35.44 | 38.46 | Negative | Negative | 1 | 32.73 | 35.05 |
| POOL 3 | 22.02 | 22.64 | 22.26 | 22.99 | 2 | 19.02 | 19.96 |
| POOL 4 | 16.66 | 16.82 | 16.84 | 16.78 | 1 | 13.54 | 13.22 |
| POOL 5 | 20.57 | 20.98 | 20.96 | 21.25 | 1 | 17.39 | 17.85 |
| POOL 6 | Negative | 27.72 | Negative | Negative | Inconclusive | Negative | 37.14 |
| POOL 7 | 24.13 | 25.31 | 27.36 | 29.06 | 1 | 20.37 | 20.23 |
| POOL 8 | 35.41 | 38.97 | Negative | 38.42 | 1 | 33,79 | 36.28 |
| POOL 9 | 19.35 | 21.18 | 19.81 | 21.53 | 2 | 16.06 | 17.39 |
| POOL 10 | 21.04 | 21.94 | 20.69 | 20.59 | 2 | 18.17 | 18.82 |
| POOL 11 | 21.61 | 23.29 | 21.48 | 23.42 | 5 | 19.12 | 18.91 |
| POOL 12 | 31.21 | 33.27 | 30.41 | 33.23 | 2 | 27.07 | 29.23 |
| POOL 13 | 20.54 | 20.32 | 24.48 | 25.12 | 4 | 20.89 | 20.55 |
| POOL 14 | 24.08 | 27.24 | 20.79 | 22.36 | 3 | 18.23 | 19.39 |
| POOL 15 | 17.00 | 18.21 | 16.97 | 18.18 | 3 | 13.55 | 15.01 |
| POOL 16 | 28.89 | 31.42 | 28.81 | 31.23 | 2 | 25.47 | 27.17 |
| POOL 17 | 20.50 | 21.41 | 20.96 | 21.96 | 3 | 17.97 | 18.74 |
| POOL 18 | 27.57 | 29.11 | 26.54 | 28.7 | 2 | 24.05 | 25.12 |
| POOL 19 | 21.34 | 22.06 | 21.41 | 22.40 | 1 | 18.13 | 18.59 |
| POOL 20 | 32.61 | 34.17 | 32.54 | 39.07 | 1 | 29.33 | 30.51 |
| POOL 21 | 28.02 | 29.26 | 26.99 | 27.26 | 2 | 23.68 | 24.26 |
| POOL 22 | 19.23 | 19.73 | 19.31 | 19.76 | 1 | 16.11 | 16.74 |

\*Protocol A: Pooling of clinical samples prior to perform RNA extraction.
\*\*Protocol B: Pooling samples after individual RNA extraction.

### Results

There was 100% and 91% of agreement when the results from individual and pooled samples with Protocol A (22 of 22) and Protocol B (20 of 22), respectively, were compared. The Cts of the individual samples with positive results ranged from 13.22 to 37.14, for both N1 and N2 targets. The Cts of the two individual samples with divergent results in Protocol B were >32.73 – Table 1. RNAse P used as a quality control presented very satisfactory results (Cts between 26 to 32).

It was possible to observe that Cts (both targets) of pooled samples in Protocol A with positive results in the RT-qPCR tended to be 3 units higher in average when compared to Cts of the individual tested positive samples (Table 1). This increase was also observed in pools of the Protocol B for the N1 target but the Ct of N2 target presented a difference of 4 units in average compared to the individually tested samples. Noteworthy, in pools with more than one positive sample, Ct of the pool was similar to the lowest Ct obtained in the individual analyses of the samples included in the pool (Table 1).

## Discussion

A rapid and reliable laboratory diagnosis of SARS-CoV-2 infection plays a crucial role for public health interventions related to the new coronavirus pandemic (3). RT-qPCR detection of the virus by oro/nasopharyngeal swabbing is characterized by good sensitivity and high specificity and has been regarded as the "gold standard” for the virus genome detection (4). However, it is still a semi-automated strategy which includes several sequential analytical steps and is associated with significant costs. Sample pooling is a well known approach to optimize time and costs of massive screening strategies for infectious diseases and has been reported as a reliable technique for Hepatitis B, Hepatitis C and Human Immunodeficiency Virus in blood banks (5).

In this study, we demonstrate that RT-qPCR with primers for the nucleocapsid of SARS-CoV-2 using pooled samples (10/pool) is able to provide a correct diagnosis even when only one individual sample is positive in the pool. In fact, the pool RT-qPCR analysis identified a positive sample (Protocol A) even when the single sample analysis had a later Ct (Ct > 32.73). As previously described, pools with a high number of samples (30 samples) may present false-negative results in the RT-qPCR for SARS-CoV-2 especially when a positive sample has a later Ct (6). Therefore, according to our experience, we suggest to make pools with a maximum of 10 samples/pool as this approach increases significantly the diagnostic capacity of the labs without losing quality.

However, the size of the pool can accommodate different infection scenarios and it can be adequate if necessary, according to the institution necessity as suggested by Lohse et al (6). It has to be considered that the use of pools would present best performance whether the population in analysis is expected to present low incidence of the COVID-19 such as people without symptoms, for example.

The best performance of Protocol A in relation to Protocol B in our study is probably related to a larger volume of each individual clinical sample added to the pool (100μL of clinical sample). Furthermore, Protocol A is operationally straightforward and less time consuming as it requires only one RNA extraction. Moreover, the only pool which contained an individual sample with inconclusive result presented inconclusive result only using the Protocol A.

According to Buckingham et al (7), the absolute Ct comparison is only meaningful when comparing experiments using the same reaction conditions. In this study, the individual and pooled samples were tested together in the same experimental run, and using the same reagents. Considering the Ct differences between a positive individual sample and the same sample tested in a pool, we observed an average variation of 3 units of Ct (Protocol A). According to the literature, using the same sample in a 10-fold dilution series, the Ct values differ by ~3.3 units (7). These data corroborate with the Ct variation found in this study between the result of the RT-qPCR for SARS-CoV-2 of the individual sample and of this sample in a pool with nine others. We also found that pools with more than one positive sample present Cts more related to the lower Ct of the sample tested individually.

Finally, the results of this study indicate that it is possible to obtain positive results of RT-qPCR for SARS-CoV2 when combining 10 clinical samples in one pool (Protocol A). The pooling of clinical samples would increase significantly test capacity of the laboratories. The use of pools would be much more effective to test clinical samples in scenarios of low prevalence of SARS-CoV-2 as a positive result of the pool will require that all samples be tested individually. In conclusion, to test populations in a large scale, the use of pools is a very useful strategy as an epidemiological screening method.

## Data Availability

The authors confirm that the data supporting the findings of this study are available within the article.

## Acknowledgments

This study received a grant of “Fundação de Apoio a Pesquisa e Ensino do rio Grande do Sul (FAPERGS)” (#20-2551-0000265-9) as well as of “Fundo de Incentivo a Pesquisa e Eventos do Hospital de Clínicas de Porto Alegre (FIPE/HCPA) – (#20200163).

